# The Relative Contributions of Arousal Presence and Arousal Intensity to Post-Respiratory-Event Ventilation in Obstructive Sleep Apnea

**DOI:** 10.1101/2025.08.30.25334191

**Authors:** Junyan Zhang, Dwayne L Mann, Timo Leppänen, Ali Azarbarzin, Scott A Sands, Juha Töyräs, Philip I Terrill

**Affiliations:** School of Electrical Engineering and Computer Science, The University of Queensland, Brisbane, Queensland, Australia; Department of Technical Physics, University of Eastern Finland, Kuopio, Finland; Clinical Research Center, Kuopio University Hospital, Kuopio, Finland; Brigham and Women’s Hospital and Harvard Medical School, Boston, Massachusetts, USA; Science Service Center, Kuopio University Hospital, Kuopio, Finland

**Author notes:** Corresponding Author: Junyan Zhang, MEng, Beng, School of Electrical Engineering and Computer Science The University of Queensland, Brisbane, Australia. **Author Contributions:** Conception and Study design: T.L., S.A.S, J.T., P.I.T.; Algorithm development: J.Z., A.A., S.A.S., D.L.M., P.I.T.; Data analysis: J.Z. All authors interpreted data, edited manuscripts for intellectual content and approved the manuscript. **Data availability statement:** The MESA data set is publicly available from NSRR repository. The Physiological Study data set used and analyzed in the current study is available from the corresponding author upon reasonable request. **Conflict of Interest:** No conflict of interest to declare.

**Keywords:** Airway collapsibility, Arousal threshold, Endotypes, Obstructive Sleep Apnea, Respiratory physiology, Sleep-disorder breathing

## Abstract

**Study Objectives:** Obstructive sleep apnea (OSA) exhibits cyclical patterns of respiratory events followed by hyperpneic breaths which frequently coincide with arousals. While this hyperventilation partially results from accumulated respiratory stimuli, arousal (i.e., its presence and intensity) appears to contribute independently. As such, this study aims to quantify the relative contributions of arousal presence and intensity to post-event ventilation independent of chemoreflex-driven responses.

**Methods:** This study utilized two retrospective polysomnography (PSG) data sets, comprising a community-based (Multi-Ethnic Study of Atherosclerosis, MESA, N=1781) and the Physiological Study data set (N=67). Arousal presence (0/1), arousal intensity (range 0-9), and post-event ventilation were calculated for each respiratory obstructive event for all participants using computational models. Mixed-effects linear models were used to examine the association between arousal presence and intensity, and post-event ventilation at both inter-event and inter-participant levels.

**Results:** At the inter-event level, arousal presence at respiratory event termination increased ventilation by 17-30%Eupnea (increase above eupneic baseline) in both data sets, where each step increment in arousal intensity contributed an additional 1-4%Eupnea increase in ventilation. At the inter-participant level, each step increment of overnight mean arousal intensity was associated with 2-4%Eupnea increase in ventilatory response to arousal in both data sets.

**Conclusions:** Our findings demonstrate that arousal presence plays an essential role in elevating post-event hyperventilation in OSA, with arousal intensity exerting modest influence per increment, but substantial cumulative effects at moderate-high levels. These results, consistent across both data sets, suggest a distinct mechanistic pathway to post-event hyperventilation beyond chemoreflex stimulation.

## Introduction

Obstructive sleep apnea (OSA) is associated with daytime symptoms such as impaired vigilance, and excessive daytime sleepiness in addition to long-term health implications, including cardiovascular disease (1–5). As such, understanding the underlying pathophysiological relationships leading to the manifestation of OSA is an essential step for developing improved diagnostic, prognostic and treatment strategies. In particular, the relationship between arousals associated with obstructive respiratory event termination (‘respiratory arousal’) and the associated period of hyperventilation is not yet fully understood.

A characteristic pattern of moderate-to-severe OSA is the cyclic ventilatory behavior, whereby the termination of obstructive apneas and hypopneas is followed by 1-3 hyperpneic breaths, which are often (but not always) temporally associated with a respiratory arousal. This cycle may then repeat with the onset of the next apnea or hypopnea (6–8). However, the mechanistic role of arousal in this pattern of behavior is complex and not yet fully understood (6–9). In particular, while post-event hyperpneic breaths are partially explained by chemoreceptor feedback mechanisms and are present in the absence of arousal, there is evidence that the presence of arousal itself further contributes to greater ventilation during these breaths. This arousal-mediated additional ventilation is referred to as the ventilatory response to arousal (VRA) (6, 8, 10), and varies significantly both within and between patients (10, 11). There is mixed evidence whether this intermittent transient response yields deleterious effects on ventilatory system stability or is a protective mechanism that improves airway patency (12, 13).

It is also recognized that there is variability in the characteristics (amplitude and time-frequency) of arousals, and this has been described as the “arousal intensity”, and can be quantified using a validated computational algorithm (14–17). Previously, Amatoury et al. (18) investigated the relationship between the mean overnight post-event ventilation and the mean overnight arousal intensity using a CPAP manipulation protocol. They concluded that an individual’s average arousal intensity is a distinct trait that is not associated with the pre-arousal respiratory stimulus, including arousal threshold, minute ventilation, and change of epiglottic pressure. However, the CPAP manipulation based protocol hinders the comparison between the relative effects of arousal presence and arousal intensity, and the investigation of the within-subject relationship between arousal intensity and post-event ventilation. CPAP manipulation induced respiratory events and arousals may also exhibit behavior distinct from naturalistic sleep.

As such, in this study we hypothesize that the variation of post respiratory event ventilatory response is associated with arousal presence (compared with non-arousal) and arousal intensity independent of chemoreflex related stimulus. This study investigates this association both within a participant (i.e. on an inter-event level), and as a trait between participants. We address this hypothesis with two complementary analyses in two retrospective data sets of polysomnograms (a large community cohort and a smaller cohort study as part of a physiological study). First, we examine the association between post-event ventilation and arousal intensity. Using mixed effect statistical models, we examine the relative effect size of arousal presence (vs. non-arousal event) and arousal intensity on an inter-event level; and test whether these effects are independent of respiratory event severity (as a proxy of chemoreflex stimulatory effects) as quantified by the event-specific ventilatory burden of the associated respiratory event. Second, we apply our computational modelling-based approach to estimate the mean overnight VRA (distinct from chemoreflex); and use linear regression models to examine the inter-participant relationship between mean overnight VRA and mean overnight arousal intensity.

## Method

### Participants and Data

#### Data Set 1: Multi-Ethnic Study of Atherosclerosis (MESA)

The MESA is a United States-based study to assess risk factors for cardiovascular disease in four different ethnic groups consisting of 6814 men and women (19, 20). A subset of participants (N=2055) conducted a successful in-home polysomnography (PSG) study with the Compumedics Somte System (Compumedics Ltd., Abbostville, Australia) and have valid PSG recordings. The present analysis utilized the airflow signal from PSG which was recorded via nasal pressure transducer. All PSGs were scored according to American Academy of Sleep Medicine (AASM) 2012 criteria (apneas detected with >90% of flow reduction with no oxygen desaturation or arousal requirements; hypopneas detected with > 30% flow reduction, and to be associated with≥ 3% oxygen desaturation or with arousal) (21).

#### Data Set 2: Physiological Study

A cohort of 77 participants with suspected OSA was previously recruited for a physiological sleep study. The recruited patients attended a single overnight in-laboratory PSG with additional instrumentation including an oronasal mask with pneumotachograph, and intra-esophageal catheter. This study was approved by the Mass General Brigham Institutional Review Board (Identifier: 2019P000666). The additional physiological measurements were used as part of a detailed study addressing a broader range of research questions (22–25). For our analysis, we used airflow recorded via pneumotachograph and diaphragmatic electromyograph (Edi) recorded via the intra-esophageal catheter as the reference for ventilatory drive. PSGs were scored according to AASM 2012 criteria. As originally scored, arousals in the Physiological Study were carefully scored and had no upper limit on duration to ensure an accurate representation of arousal and wake events (21). We employed a 15-second limit on durations for the current study to satisfy the requirements for arousal intensity calculation (14).

### Participant Selection and Exclusion

Participants in the two data sets were included in analyses if they had an apnea hypopnea index (AHI) ≥5 events/h; and signal quality and scoring of respiratory events, arousal events and sleep stages was adequate for analysis. From the initial 2055 PSG recordings in MESA data set, 214 participants had an AHI<5 events/h, 19 participants had incomplete scorings, and a further 52 participants had inadequate signal quality, leaving 1781 participants for the analysis (Figure 1). In the Physiological Study data set, 9 participants had the AHI<5 events/h, and 1 participant had inadequate signal quality during respiratory events, leaving 67 participants for this study.

**Figure 1.**
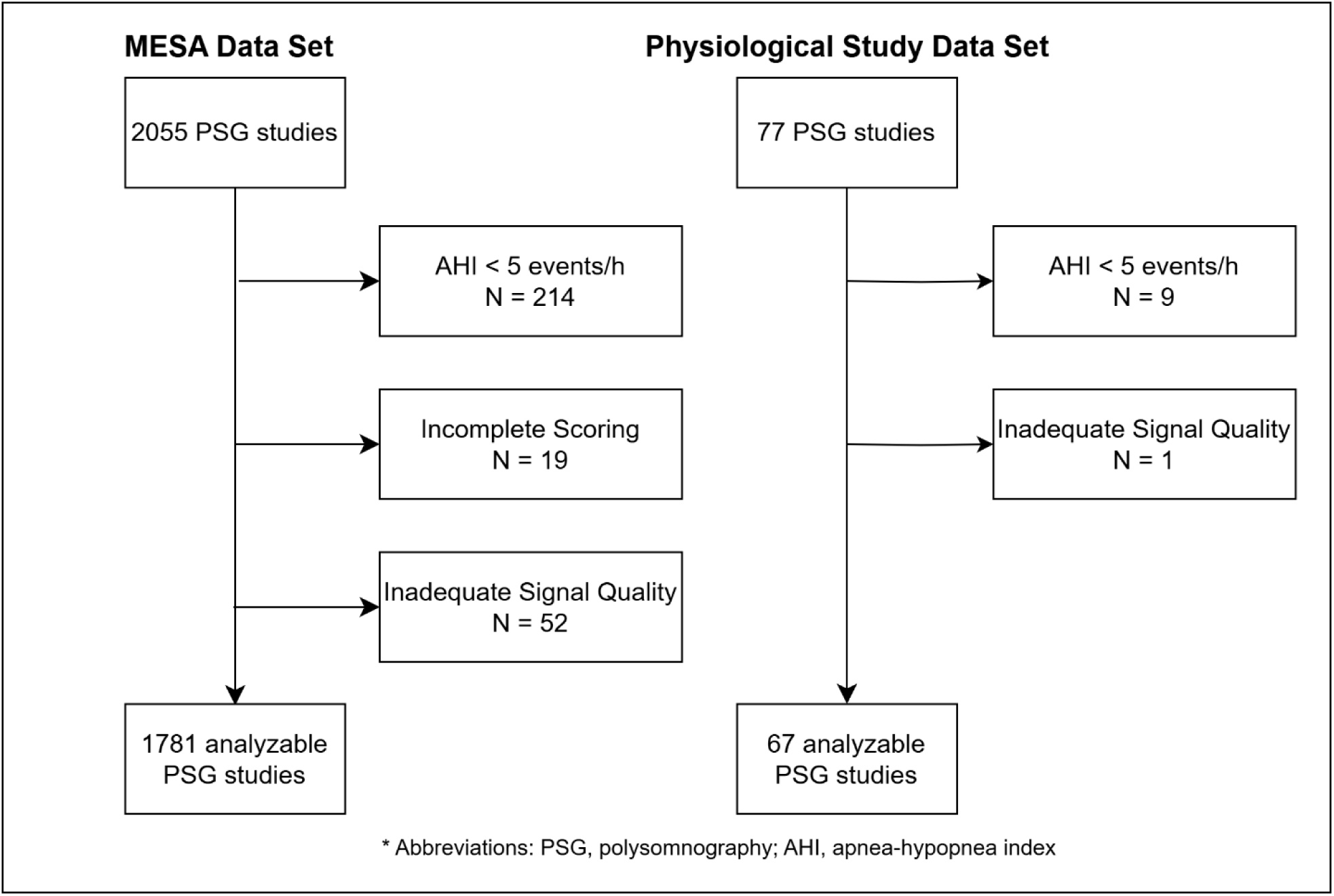
Flowchart of applying exclusion criteria to both data sets.

### Signals analysis and processing

#### Quantification of arousal presence and arousal intensity

Arousals were characterized through two primary variables: *a) Arousal presence*: A binary categorical variable indicating whether the respiratory obstructive event terminated with or without an arousal (0 = no-arousal event, 1 = arousal event); and *b) Arousal Intensity*: which was measured on a discrete scale between 0 and 9 (9 being the most intense arousal), using a validated automated wavelet transformation/time-frequency analysis based machine learning algorithm (14, 26). Arousals whose onset occurred within +/- three seconds of the termination of a scored hypopnea/apnea event were defined as respiratory arousals (27–29). All other arousals were classified as spontaneous arousals and were excluded from the current study. Arousals and respiratory events within REM sleep were also excluded. The mean arousal intensity for each subject was calculated using all valid respiratory arousal events.

### Quantifying Obstruction Severity and Post-event Ventilation

The breath-by-breath ventilation (**v**_E_) was derived from the nasal pressure signal (MESA data set) or pneumotachograph signal (Physiological Study data set) and presented as percentage of eupneic ventilation (%Eupnea), which was defined as the mean ventilation per each 7-minute analysis window.

Post event ventilation and Obstruction severity for an event were quantified as follows (**Figure 2**): *a) post-event ventilation*: defined as the mean ventilation of the first two breaths immediately following event termination; *b) Ventilatory burden*: calculated on an event-by-event basis and defined as the cumulative loss in ventilation in each respiratory obstructive event (30). The event-specific ventilatory burden can be calculated as:

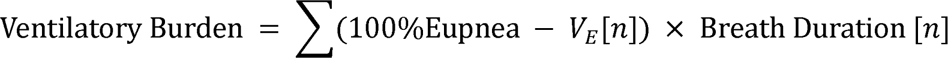

where *n* is the n^th^ breath of the given scored obstructive event. The event-specific ventilatory burden is expressed in %Eupnea⋅sec, whereas post-event ventilation is expressed in %Eupnea.

**Figure 2.**
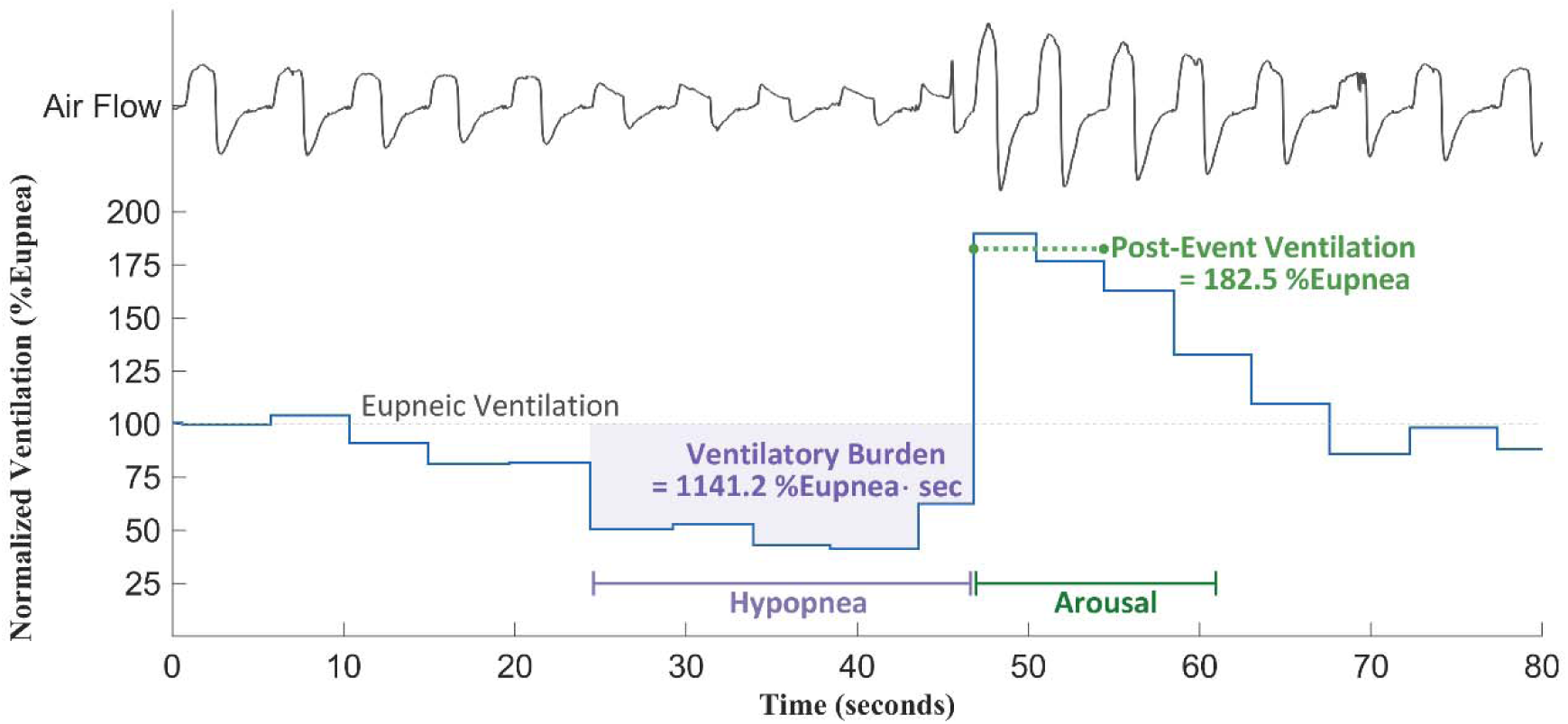
Illustrating the Post-Event Ventilation and Event-Specific Ventilatory Burden in an example hypopnea event followed by an arousal.

### Computational model-based estimation of chemoreflex drive and VRA

Breath-by-breath ventilatory drive was estimated using a computational method (22, 31); which fits a first-order model of the chemoreflex feedback system in a 7-minute overlapping sliding window from the airflow signal. The model-estimated ventilatory drive is comprised of two components: 1) the chemoreflex drive (**v**_Chem_) and 2) the additional arousal-invoked drive that is only present during arousals (i.e., VRA), both expressed in %Eupnea (31). The overnight mean VRA is computed by averaging the window estimates of VRA across the night for each participant. We also computed overnight endotypes including loop gain, muscle compensation, collapsibility and arousal threshold, and evaluated their associations with overnight mean arousal intensity.

To provide an estimate of chemoreflex ventilatory drive at the termination of each respiratory event in inter-event analysis, we also extend this model to calculate the chemical drive at the last breath of each obstruction event. This is equivalent to the event-specific arousal threshold for events terminated with an arousal. For the Physiological Study Data Set only, the Edi signal is used as the reference ventilatory drive at the last breath of each obstruction event and assuming the mean of the ventilatory drive is 100%Eupnea in the same 7-minute window.

### Data Analysis

#### The event-level relationship between arousal presence, arousal intensity and post-event ventilation

The ventilation (**v**_E_) during the respiratory event and the post-event periods was first assessed using an ensemble-averaging method. Within each subject, all the scored respiratory obstructive events were stratified into three categories based upon arousal presence and intensity: high-intensity (arousal intensity ≥5), low-intensity (arousal intensity <5), and no arousal. Arousals were stratified into a binary classification of high-intensity and low-intensity for investigating the ventilatory characteristics difference between high- and low-intensity events. In total, 40360 and 2132 high-intensity events; 40446 and 1784 low-intensity events; and 79264 and 1136 obstructive events that were not terminated with arousal were identified and analyzed in the MESA data set and Physiological Study data set respectively. Each respiratory obstructive event was then aligned at the termination breath (i.e. the last breath within which the event is present, called Breath 0 herein) and truncated to 3 breaths prior and after Breath 0. Ventilation for each category was taken as the average for each participant (participant level average). Then the overall average was taken by averaging all the participant-level average for each category. Wilcoxon signed-rank tests were used to evaluate any statistically significant differences between 1) high-intensity vs low-intensity and 2) low-intensity vs no-arousal for each breath and 3) high-intensity vs. no-arousal for each breath.

A linear mixed-effect model regression analysis was employed to model the relationship between post-event ventilation (the dependent variable) with arousal presence and arousal intensity (independent variables) included on an event-by-event basis. Participant was included as a random intercept, while arousal presence and arousal intensity were both included as fixed-effects. To effectively capture the relative contributions of arousal presence with the addition of arousal intensity, arousal presence was coded as a binary categorical fixed-effect variable (0 for no-arousal; 1 for arousal), and arousal intensity (range 0-9) was coded as an interaction term with arousal presence (i.e. Model 1: post-event ventilation = arousal presence + arousal intensity.arousal presence + random intercept) to utilize the entire set of events and preserve the scale of arousal intensity. This primary model (Model 1) was subsequently adjusted for event severity with event-specific ventilatory burden (Model 1a), and the chemical drive (estimated) at the last breath of obstruction event, Model 1b). For the Physiological Study data set only, the Model 1c is included to adjust for the ventilatory drive at the last breath of obstruction measure with Edi.

### The relationship between overnight average arousal intensity and ventilatory response to arousal

Bivariate linear regression analyses were performed to assess the inter-participant association between the overnight mean VRA (the dependent variable) and the overnight mean arousal intensity. Subsequently, models were adjusted for the participant demographics (age, sex and body mass index (BMI)) and OSA severity (AHI).

### Statistics

All analysis were performed using MATLAB (2022b, The MathWorks, Natick, MA, US). A p-value of 0.05 was considered the limit of statistical significance. All variables’ normality was tested with the Kolmogorov-Smirnov test. All mixed-effects models were examined for normality and homoscedasticity of residuals.

## Result

### Participant Characteristics

Table 1 summarizes the key demographic and OSA characteristics of both cohorts. The MESA community cohort exhibited a milder OSA severity and BMI values compared to the Physiological Study’s participants.

**Table 1.**
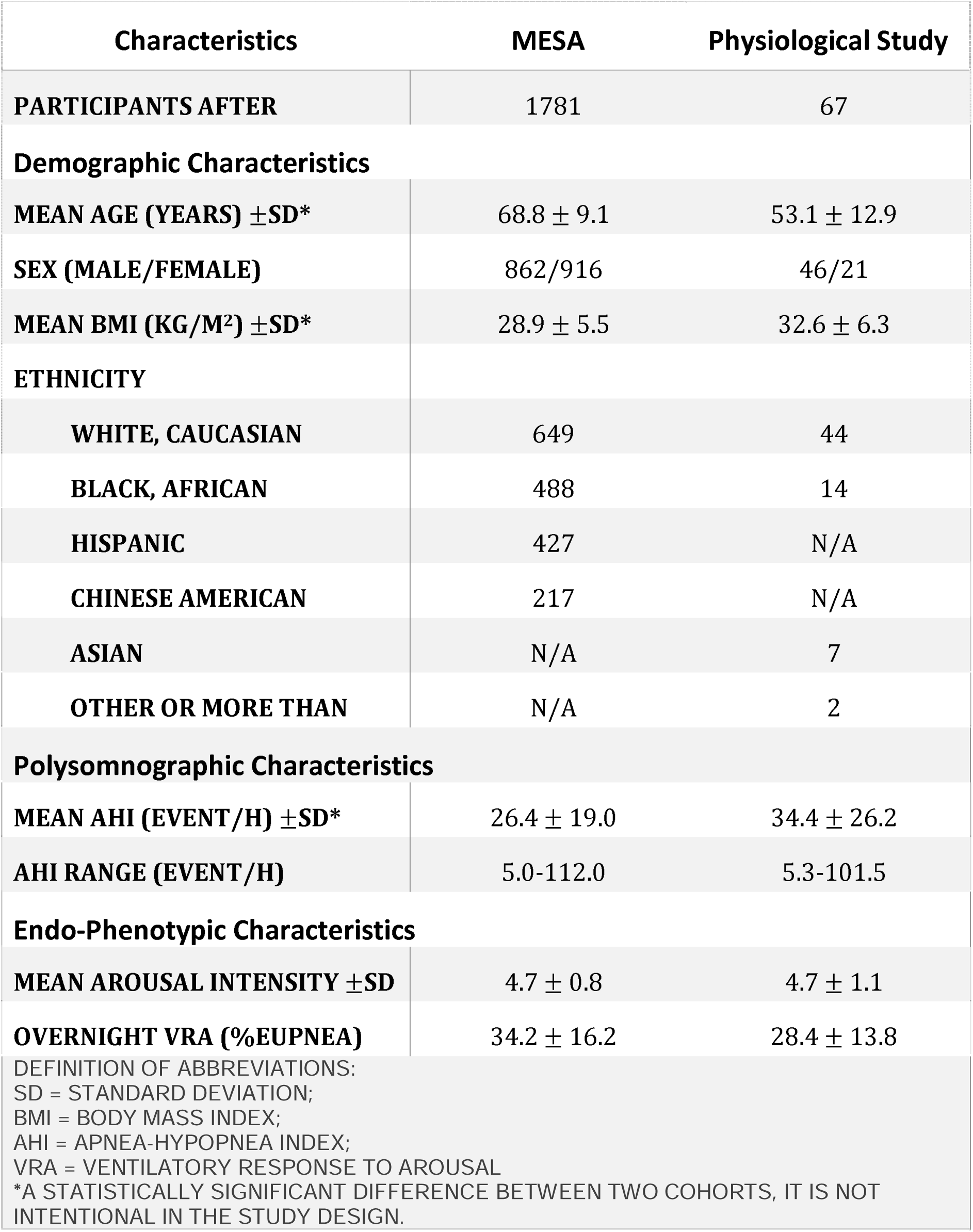
Participant Demographics, Endo-phenotypic Characteristics.

### The event-level relationship between arousal presence, arousal intensity and post-event ventilation

In the MESA data set, high-intensity arousals were associated with a statistically significant increase in **v**_E_ compared to low-intensity arousals at Breath 1 (high-vs. low-intensity vs. non-arousal: 175.5±51.0 vs.167.5±47.6 vs. 121.4±20.3 %Eupnea, mean ± SD; **Figure 3A**) and Breath 2 (162.9±47.1 vs. 152.9±40.5 vs. 118.3±19.0 %Eupnea). **v**_E_ of high- and Low-intensity events were significantly elevated compared to non-arousal events at both breaths. The Physiological Study data set showed similar trends, though differences between high- and low-intensity events were not statistically significant (Breath 1 high-vs low-intensity vs non-arousal: 179.1±46.3 vs. 173.5±41.1 vs. 140.1±33.7 %Eupnea, **Figure 3B**; Breath 2: 174.0±43.9 vs. 163.8±37.7 vs. 123.5±41.0 %Eupnea). However, **v**_E_ of high- and low-intensity events remained significantly elevated compared to non-arousal events at Breath 1 and 2. These results were similar when expressed as a relative change in ventilation compared with the last breath of the respiratory obstructive event (Supplementary Figure E1)

**Figure 3.**
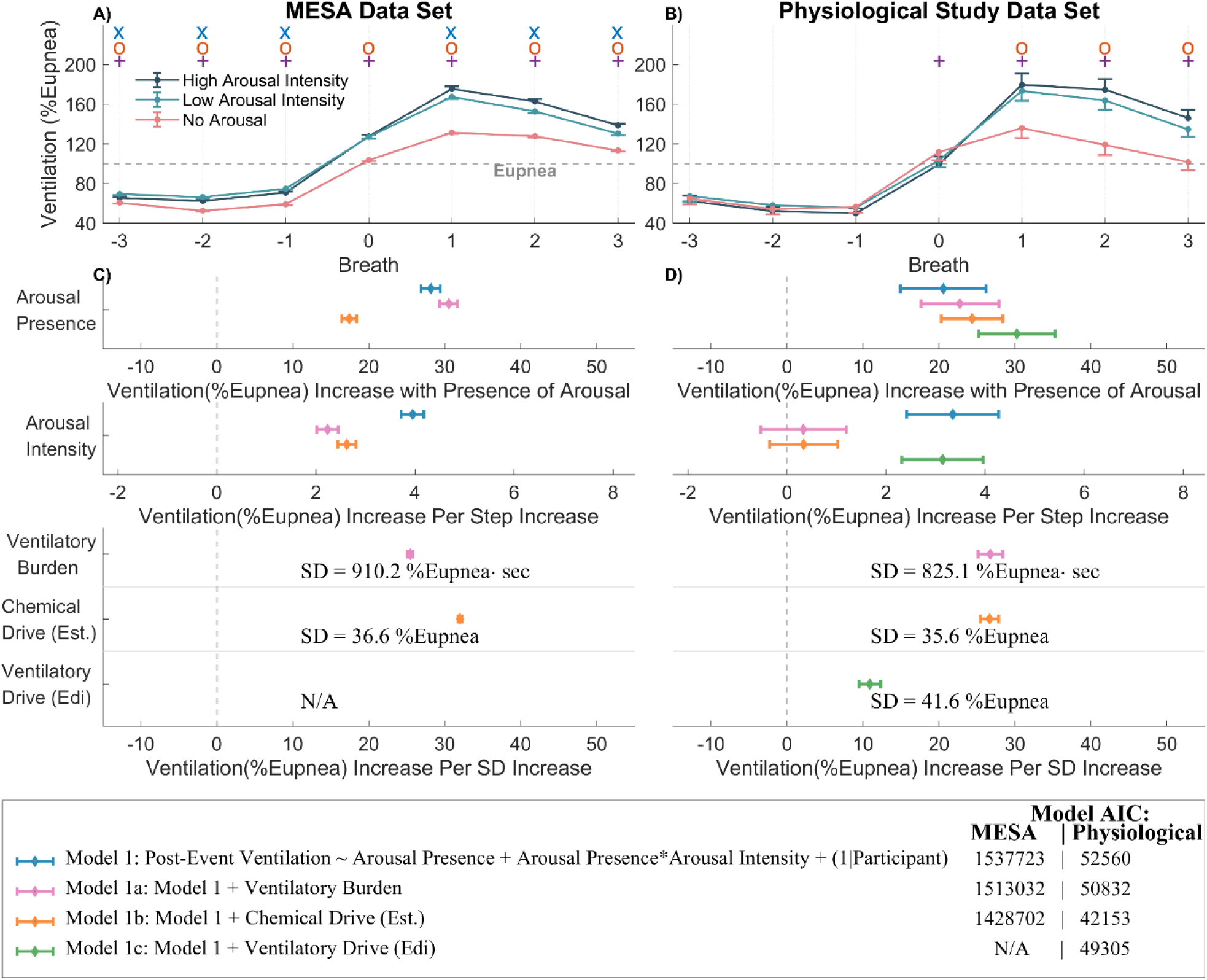
A) and B): Ventilation normalized against eupneic ventilation at the termination of respiratory events for low (0 ≤ Arousal intensity < 5) and high (Arousal intensity ≥ 5) arousal intensity events, and obstructive events not terminated with arousals for the three breaths prior (breath −3 to −1), the breath during arousal onset or event termination (breath 0) and three breaths following (breath 1 to 3) arousal or event termination. “x” represents p-value <0.05 between high and low arousal intensity event at the respective breath; “o” represents p-value < 0.05 between the low-intensity event and no-arousal event at the respective breath; “+” represents p-value < 0.05 between the event with high-intensity arousal and no-arousal at the respective breath. C) - D): Forest plots showing the association between the increase in predictor variables and the increase in %Eupnea of post-event ventilation (the dependent variable). Each color represents a separate mixed-effects linear model. Model 1 is a multivariate model that includes both arousal presence and arousal intensity. Model 1a, 1b and 1c (for Physiological Study only) are adjusted for obstruction severity with different obstruction severity metrics. ”Ventilatory Burden”: is event-specific and calculated at an event-by-event basis. “Chemical Drive (Est.)”: The estimated chemical drive at Breath −1, it is equivalent to the event-specific arousal threshold when event is terminated with an arousal. “Ventilatory Drive (Edi)”: The ventilatory drive measured with intra-esophageal diaphragmatic electromyograph at Breath −1. “IIl” represents the coefficient estimate and the horizontal error bar represents the 95% CIs.

In both cohorts, the mixed-effects models (**Figure 3, C to D**) revealed that arousal presence had a substantial impact on post-event ventilation. Model 1 shows that arousal presence was associated with a 28.2[95%CI, 26.9, 29.4] %Eupnea increase in post-event ventilation in MESA and a 20.6[14.9, 26.2] %Eupnea increase in the Physiological Study data set respectively. A one increment change in the arousal intensity yielded a more modest increase in post-event ventilation, being 4.0[3.7, 4.2] and 3.3[2.4, 4.3] %Eupnea increase respectively. Arousal presence remained as a statistically significant predictor in both cohorts after adjusting for obstruction severity with different metrics (Model 1a, 1b). While showing a more modest tend, arousal intensity remain statistical significance in the Physiological Study data set after adjusting for ventilatory drive (Model 1c) but was not statistically significant after adjusting for event-specific ventilatory burden or chemical drive (estimated) as additional fixed effects. These results were not sensitive to the temporal threshold for linking an arousal to a respiratory event (see Supplementary Figure E2).

### The relationship between overnight average arousal intensity and ventilatory response to arousal

While inter-subject variability was substantial, the result suggests there was a weak positive between-subject association between mean arousal intensity and overnight mean VRA in both data sets (**Figure 4 A-B**). This association is statistically significant in the MESA data set but borderline in the smaller Physiology Study data set. A unit increase in mean arousal intensity is associated with a 2-4%Eupnea increase in mean overnight VRA in both data sets, and no meaningful changes after adjusting for participants’ demographics and OSA severity were observed (**Figure 4 C-D**).

**Figure 4.**
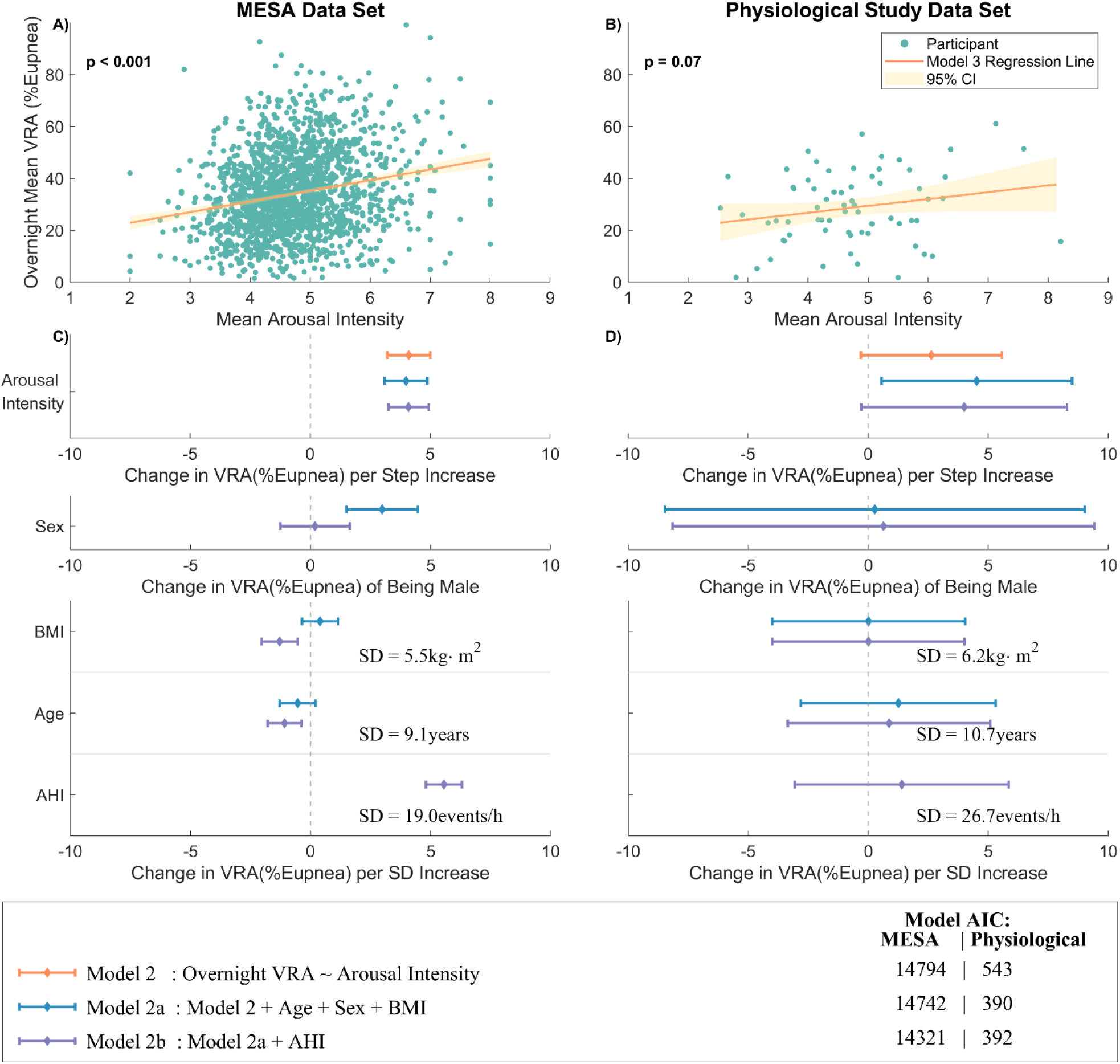
A) and B) Scatter plots showing overnight mean VRA is associated with mean arousal intensity. Each dot represents one participant. The regression line represents the unadjusted model (Model 2). C) – C): Forest plot showing the association between overnight mean VRA and mean arousal intensity for both unadjusted (Model 2) and models adjusted for subject demographics and OSA severity (Model 2a and 2b). “IIl” represents the coefficient estimate and the horizontal error bar represents the 95% CIs.

In secondary analysis, the estimated event-specific VRA showed a weak statistically significant association with arousal intensity in the MESA data set level(1.4 [95%CI: 1.3, 1.5] %Eupnea per step increment in arousal intensity). The association was not statistically significant in the Physiological Study data set (effect size: 0.4 [-0.1,0.8] %Eupnea per step).

## Discussion

The objective of this study was to investigate and quantify the relationship between arousal presence and intensity, and the associated post-respiratory event ventilation. This was investigated in two different retrospective cohorts: a United States-based community cohort conducted as an in-home PSG; and a cohort of suspected or diagnosed OSA patients who were recruited for an in-laboratory physiological study. Our inter-event analysis indicates that both arousal presence (vs. non-arousal) and arousal intensity were associated with post-event ventilation independent of event severity and estimates of chemo-reflex stimuli. While arousal intensity was showing a modest per-increment effect size, it had a similar effect size as arousal presence across its full 0-9 scale. Using a computational physiological model-based estimate, we identified that whilst there was substantial inter-participant variability, the mean overnight VRA was associated with the mean overnight arousal intensity at an inter-participant level. Overall, our results support the hypothesis that variations in both arousal presence and arousal intensity contribute to the variation in post-respiratory event independent of chemoreflex stimuli.

### The relationship between post respiratory event ventilation and arousal presence and intensity

There has been a substantial body of previous work which has investigated the relationship between post-respiratory event ventilation and arousal. In particular, several studies have observed elevated ventilation following event termination in temporal association with arousal (vs. events terminated without arousal) (6, 8, 10, 32). However there has been contention as to whether this effect is independent of respiratory event severity (i.e. consequent chemoreflex stimuli from a prolong obstruction episode) (6, 13, 33). Eckert and Younes (33) suggested the elevated post-event ventilation observed during arousal is a result of the compounding of additional drive related to arousal and elevated chemical drive, but the relative effect size from each contributor remained insufficiently quantified (33). This work has also been extended through CPAP manipulation-based protocol, demonstrating that increased arousal intensity was associated with increasing magnitude of observed post-respiratory-event ventilation between patients (6, 18, 33). In this study, we build on these previous work to examine the relative contributions of *both* arousal presence and arousal intensity to post-respiratory event ventilation in naturalistic sleep using two broad approaches that can be applied to observed polysomnogram.

Firstly, we conducted an inter-respiratory event-based analysis comparing observed post-event ventilation between events with and without arousal, and by arousal intensity. Our results across our visualization (using ensemble average), and our unadjusted and adjusted mixed-effect statistical analysis in both the Physiological Study and the MESA data sets showed reasonable consistency: arousal presence (vs. non-arousal events) was associated with a ≈20-35%Eupnea increase in ventilation; with arousal intensity contributing ≈1-4%Eupnea additional ventilation per increment. This dose-dependent effect on ventilation with arousal intensity was also observed by an alternative approach that stratified arousal intensity into discrete bands, without using linearity as the prior assumption (See Supplementary Figure E3). As expected, confidence intervals were smaller in the much larger MESA data set. The ensemble average showed only small differences in **v**_E_ between the high- and low-intensity arousals (statistically significant only in the MESA data set), likely to reflect the distribution of arousal intensity clustered between 3-6 (i.e. near the threshold of 5 for high/low-intensity arousals). We also noticed the non-arousal events in MESA data set have lower **v**_E_ than arousal events during obstruction phase, which can be a selection bias in the AASM scoring rules that requires hypopnea to be associated with oxygen desaturation or arousal event (See Supplementary Figure E4). To account for this, statistical models were adjusted by estimates of respiratory event ventilation history to distinguish between post-event ventilation explained by arousal vs. chemoreflex stimuli using event-specific ventilatory burden (“lost” ventilation relative to eupnea during the event, Model 1a), the model estimated chemical drive (Model 1b), and the measured neurological drive (Model 1c, Physiological Study only) at the last breath of the event. The adjustment had little influence on the effect size of arousal presence but reduced the effect size of arousal intensity in MESA. However, in the Physiological Study, the effect size of arousal intensity reduced to ≈0.5-1%Eupnea in Model 1a and 1b, and while borderline, were not statistically significant. The differences between the data sets may reflect sample size, and possibly the different demographics (see Supplementary Figure E5 and E6).

Secondly, we applied a physiological modelling-based approach which provides an overnight mean VRA in an inter-participant analysis. While there was substantial inter-subject variability, the overnight VRA showed a trend for a weak association with the mean overnight arousal intensity (i.e. participants who tend to have a higher VRA tend to have a higher mean arousal intensity). The trends were similar in the two data sets, and were statistically significant in the larger MESA data set with an effect size of ≈4%Eupnea per arousal intensity increment which was independent of AHI and key demographics (age, sex, BMI).

There was consistency in the results between our two separate analytical approaches. In particular, the effect size of arousal intensity on VRA in inter-participant analysis was similar to the effect size calculated in inter-event analysis. Whilst methodologies differ substantially, this is consistent (albeit lower in magnitude) with results reported by Amatoury et. al (18) who showed post-event minute ventilation is ≈40%Eupnea greater in a high-intensity arousal participant group vs. a low-intensity arousal participant group. With mean arousal intensities of 7.4 vs 3.4 respectively, this ascribes roughly a ≈10%Eupnea increase per arousal intensity increment. However, the CPAP manipulation protocol used by Amatoury et. al. did not allow the comparison between the relative effect of arousal presence and arousal intensity (18).

Furthermore, in other works, arousal (regardless of the intensity) was associated with ≈30-50%Eupnea increase in ventilation using CPAP dial-down protocol (6, 8), which aligns with our analysis showing an ≈20-35%Eupnea increase in ventilation with arousal presence. Overall, our results provide convincing evidence that post-respiratory event ventilation is associated with both arousal presence and arousal intensity; and that this is likely independent of chemoreflex stimuli.

### Physiological Insights and Clinical Implications

One of the key objectives of this study was to directly compare the relative effects of arousal presence and arousal intensity on post-event ventilation. In particular, we observe that while each incremental step in arousal intensity contributes only a modest increase in post-event ventilation, the cumulative effect across the 10-level intensity scale could be substantial. For instance, in the MESA cohort (Model 1a), arousal presence was associated with a ≈ 30.5%Eupnea increase in ventilation, and each arousal intensity increment contributes an additional ≈ 2.2%Eupnea. As such, an arousal with an intensity of 5 (population mean = 4.7) corresponds to a 41.5 %Eupnea (as 30.5%Eupnea + (5 × 2.2%Eupnea)) increase in ventilation (relative to non-arousal). Previous studies have shown the complex role of arousal in OSA pathophysiology. While arousal may serve as protective mechanism in restoring airway patency, it could also lead to destabilization of ventilatory control and promoting subsequent respiratory obstructive events (6, 8, 13, 18). Our results add to previous data indicating this potentially ventilatory destabilizing effect appears to be increased with higher intensity arousals (6, 18), suggesting a physiological trade-off wherein the protective benefit may be partially offset by the deleterious effect on breathing patterns.

Furthermore, there has been substantial work investigating the arousal threshold as a therapeutic target with the use of sedatives (34–36). Whilst there are no obvious therapies that could specifically target arousal intensity, these results suggest that reducing arousal intensity, even without a reduction in arousal frequency, may have therapeutic benefit of reducing OSA severity.

Past literature has speculated that a series of mechanisms might have contributed to the magnitude of VRA, including 1) sudden increase in upper airway patency, 2) “startle-like” reflex due to awakening and 3) restoration of waking chemical drive (32, 34, 37–40). Our results suggest that the occurrence of arousal induces a threshold-like improvement in ventilation, with more intense arousal adding further incremental increases (6). Such increases in ventilation must be associated with some combination of improved airway patency and/or increased neurological ventilatory drive. Amatoury et. al. observed increased tensor palatini activity in association with arousal intensity, which may contribute to improved airway patency (albeit, as part of a complex response of multiple muscle groups) (18). While we did not directly measure muscle activity, in additional analysis, we estimated breath-by-breath flow-limitation (expressed as a ratio of flow to neurological drive) using a published flow-shape-based machine-learning algorithm (41), see Supplementary Figure E7. Notably, while there was a substantial improvement in flow-limitation in arousal vs. non-arousal; there was no significant difference between high- and low-intensity arousal. This suggests that while arousal is associated with substantial improvement in airway patency, increasing arousal intensity has little further benefit; and as such, increases in observed ventilation in association with arousal intensity are likely due to greater neurological ventilatory drive (above and beyond chemoreflex stimuli). Whilst not validated for this purpose, both the ensemble average of our computational model-based breath-by-breath ventilatory drive (31) and the ensemble average of the ventilatory drive derived from Edi signal support this increased ventilatory drive in low-vs. high-intensity arousals (see Supplementary Figure E8 and E9).

Model estimated OSA endotypes, including loop gain, collapsibility, muscle compensation and arousal threshold were not found to be associated with the arousal intensity in our analysis (see Supplementary Figure E10 and E11). This is consistent with Amatoury et. al.’s findings using the more invasive physiological investigation and suggests that arousal intensity does not play a mechanistic role in any of the other pathophysiological endotypical traits of OSA.

### Methodological Strengths and Limitations

This study presents several notable strengths. Firstly, in contrast to many previous studies investigating ventilatory response to arousal (6, 8, 18, 42), the current study investigated ventilatory patterns associated with arousals in a relatively naturalistic sleep environment, free from deliberate sources of arousal-inducing stimuli during periods of analysis. In addition, our findings were generally consistent with findings from previous studies employing CPAP drop manipulation and gold-standard physiological measurements (6, 8, 18) and provides external concordance to these previous data. Secondly, our study encompassed two distinct and independent OSA cohorts: one comprising a community-recruited population and another consisting of recruited OSA patients. The similarity in results observed across these cohorts provides internal concordance, thereby offering additional validation of the observed physiological phenomena. This cross-cohort consistency strengthens the generalizability of our findings and underscores the robustness of the observed arousal-related respiratory dynamics across diverse OSA populations.

However, this study has several limitations. Firstly, the physiological model calculated variables including the **v**_Chem_ and VRA were estimated with computational models. The **v**_Chem_ and VRA were estimated based on the assumptions that the physiological characteristic is relatively stationary within a short period, which may not always be satisfied in PSG data. These methods were validated in their ability to provide overnight estimates of endotypical traits rather than direct breath-by-breath estimates of drive; and due to the absence of a clear physiological gold-standard, the calculation of VRA itself has not previously been validated. However, analysis using physiological model-based analysis was consistent with our other analysis which are not making use of model derived variables (i.e., Model 1a was consistent with Model 1b). Secondly, clinical scoring of arousal and respiratory events is known to be variable and may not accurately represent the onset and duration of the arousal, therefore, there is a concern for the future translational use of the knowledge. Finally, this work focused on the cross-sectional study of arousal dynamics and its associated ventilatory characteristics in NREM sleep. Whilst we observed there is difference in the arousal and its ventilatory characteristic between NREM and REM sleep (see Supplementary Figure E12 and E13), the mechanism causing the difference is yet to be investigated. Nevertheless, the long-term effects of this arousal to ventilation association also remains unknown. Further research could employ data sets with longitudinal study design to assess the arousal-related respiratory dynamics in response to various OSA treatment interventions, including CPAP treatment and pharmaceuticals. Linking the arousal characteristics with clinical outcomes could further elucidate the clinical relevance of these findings and their importance relative to other physiological biomarkers (43, 44).

### Conclusion

Prior studies have demonstrated that the arousal presence and intensity are associated with an augmented post-respiratory event ventilatory response (6, 18). Our study extends this understanding by quantifying their relative contributions across two independent cohorts. We demonstrated that arousal exhibits threshold-like behavior on post-event ventilation: arousal presence provides the initial step increase in ventilation, with subsequent intensity contributing modest incremental effects. Furthermore, there was substantial inter-participant variability in these relationships, thus, the physiological implications of these findings varied depending on the individuals or phenotypical groupings.

## Supporting information

Supplementary Analysis and Figure

## Data Availability

The MESA data set is publicly available from NSRR repository. The Physiological Study data set used and analyzed in the current study is available from the corresponding author upon reasonable request.

