## Supplementary Analysis and Figure for "The Relative Contributions of Arousal Presence and Arousal Intensity to Post-Respiratory-Event Ventilation in Obstructive Sleep Apnea"

Online Data Supplement

### Ensemble-Average Respiratory Event Analysis (Relative to Breath -1)

To confirm that the ensemble analysis for post-event ventilation were not contingent on which breath was declared as the ‘end of respiratory event/start of arousal event’, we repeated the analysis, however setting the end of event breath as the reference breath. The relative changes of the data $(\Delta)$ to the reference breath were calculated. While anticipated subtle changes appeared, there was no meaningful change to the interpretation of results (**Figure E1**).

Figure E1

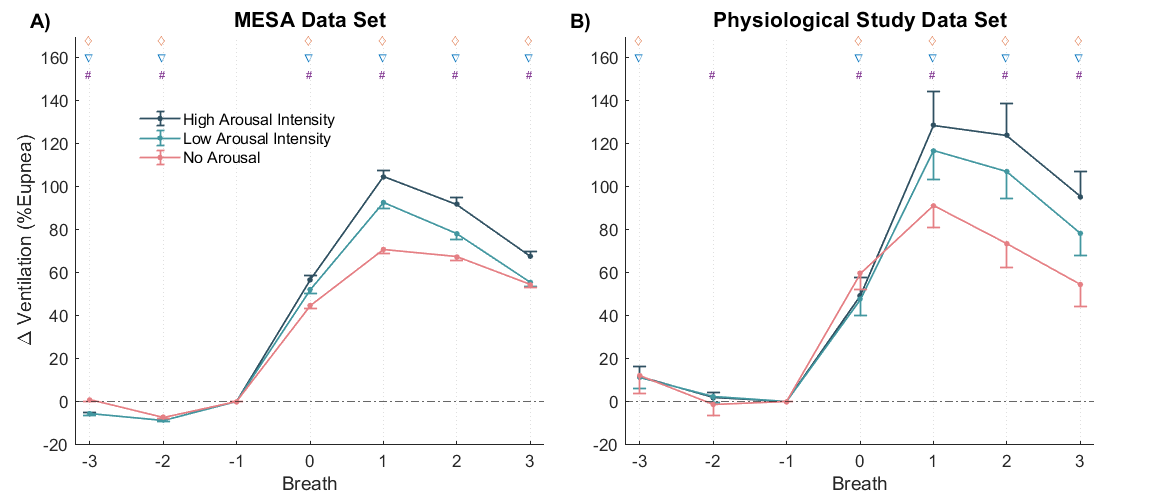

Relative change in ventilation related parameters for non-arousal, high-arousal-intensity and low-arousal-intensity events. Ventilation (Panel A and B), Chemical Drive and VRA (Panel C and D) and Flow-Drive ratio (Panel E and F), at the patient level average binned by high intensity arousal vs low intensity arousal vs event not terminated with arousal. The Breath -1 (termination threshold) were set as the baseline for each parameter. The values are presented as difference to the baseline (Δ). The marker “◊”, “▽” and “#” indicate there is a statistically significant difference between that breath to the baseline value for high-intensity, low-intensity, and events not terminated with arousal respectively.

### Sensitivity Analysis on Respiratory-related Arousal cut-off threshold

In the main analysis, the respiratory-related arousal needed to meet the following criteria: arousal onset is during a scored obstructive event or within 3 seconds from the event termination. To confirm the result from the mixed-effects models were not contingent with the 3-second cut-off point, we repeated the analysis (Model 1) with cut-off points at 1, 2, 4 and 5 seconds. While anticipated subtle changes appeared with different cut-off points, there was no meaningful change to the interpretation of results (**Figure E2**)

Figure E2

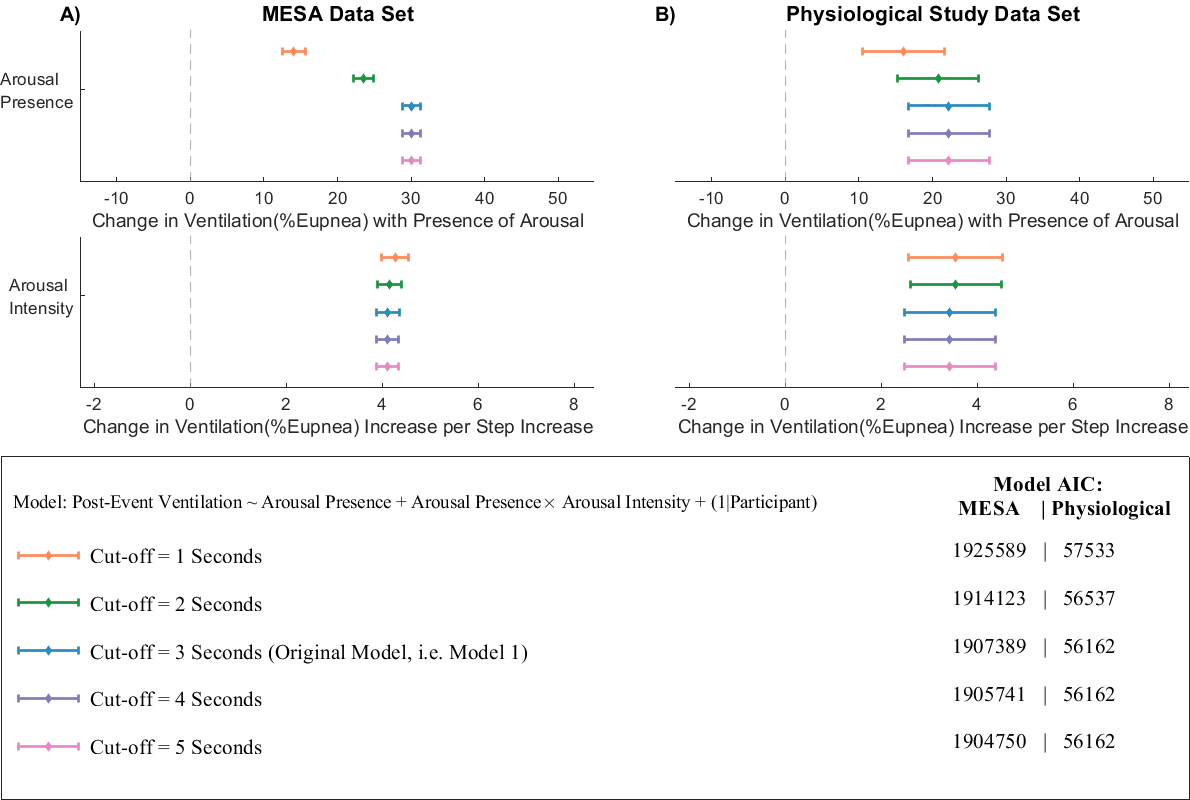

Repeated mixed-effects model analysis with different respiratory-related arousal cut-off point.

### Stratified Arousal Intensity Analysis

To address the possibility of threshold-dependent effects of arousal intensity on post-event ventilations, we conducted a complementary analysis using stratified intensity bands. This analysis specifically tests whether elevated ventilatory responses emerge only above certain arousal intensity thresholds or follow a consistent linear pattern across the full intensity spectrum.

We implemented a mixed-effects model with post-event ventilation as the dependent variable. Fixed effects included the binary arousal presence variable and three additional binary variables representing stratified arousal intensity bands: ArI34 (intensity=3-4, inclusive, same below), ArI56 (intensity=5-6), and ArI789 (intensity=7-9). Low-intensity arousals (≤2) were captured within the arousal presence effect. All binary variables were set to zero if the event was not terminated with arousal. The model adjusted for event severity by including event-specific ventilatory burden as a continuous fixed-effect variable. Participant was included as a random intercept.

**Figure E3** revealed a progressive pattern of increasing ventilatory response corresponding to increasing arousal intensity (ArI34<ArI56<ArI789) in both data set, supporting a linear rather than threshold-like relationship between arousal intensity and post-event ventilation. Notably, the gradient of increase (slope) was steeper in the MESA data set than the smaller Physiological Study data set, which is also reflected in original Model 1b.

Figure E3

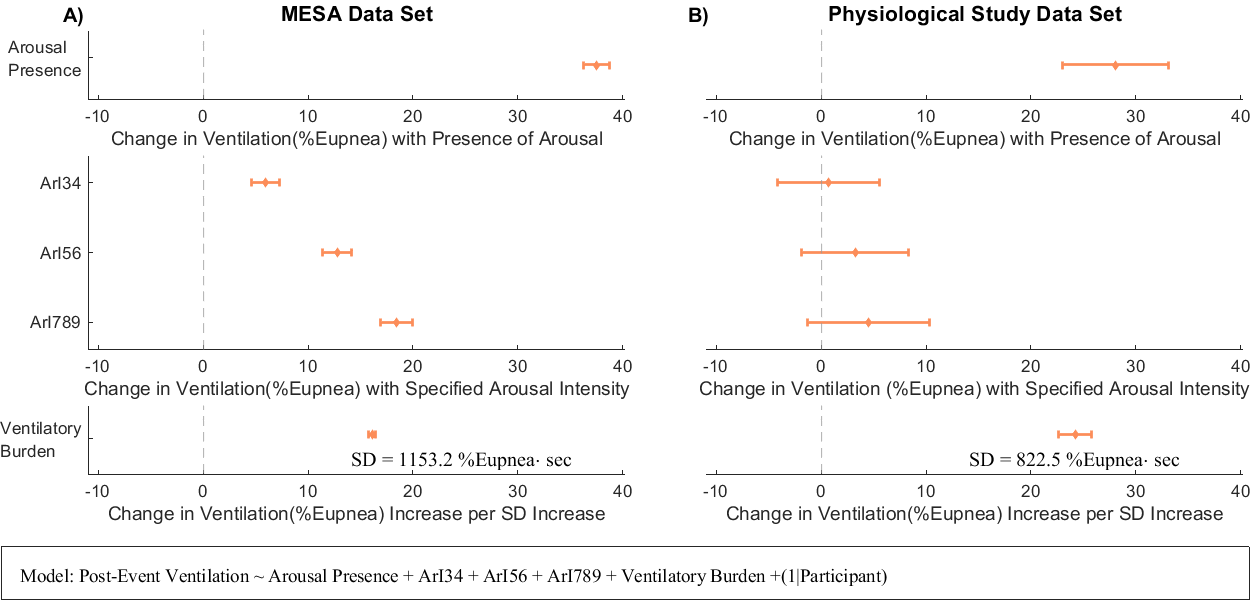

A) and B): Forest plot illustrates the effect on post-event ventilations in %eupnea. ArI34, ArI56 and ArI789 are binary variables representing stratified arousal intensity bands 3-4, 5-6 and 7-9 (e.g. If an event has an arousal intensity of 4, ArI34 = 1, ArI56=ArI789 = 0; if an event is not terminated with arousal, ArI34=ArI56=ArI789=0). “⧫” represents the coefficient estimate and the horizontal error bar represents the 95% CIs.

### Within-Participant Difference in Ventilations: Arousal vs Non-Arousal

In the ensemble average analysis within the MESA dataset, we observed that obstructive respiratory events that do not terminate with an arousal appear to have lower ventilation throughout the event compared to those events that do terminate with an arousal (**Figure E4-A**, copied from Figure 3). Initially, this may seem counterintuitive according to the traditional chemoreflex model which suggests greater degrees of obstruction incur greater chemoreflex stress that ultimately triggers an arousal (once the arousal threshold is reached). However, the physiological mechanisms that underpin obstruction and arousal are complex and are likely to have large inter-participant variability, which is being masked within the ensemble average presentation. Indeed, the median *within-participant* differences in ventilation from Breath -3 to -1 between obstructive events that terminate with an arousal versus those that do not terminate with an arousal is small but varies considerably across patients (**Figure E4-B**).

Furthermore, AASM scoring criteria requires hypopnea events to be associated with an oxygen desaturation ≥3% *or* an arousal. This creates a systematic selection bias wherein the events that terminate with an arousal can include both those with an accompanying desaturation, but importantly, also those without an accompanying desaturation. In contrast, the events that do not terminate with an arousal must have an accompanying desaturation. Consequently, the “non-arousal" group is enriched with severely obstructed events, i.e., those that elicit a desaturation. This is also supported by our analysis of flow limitation (see Supplement Figure E7) which shows that events that do not terminate with an arousal tend to have more severe flow limitation occurring during the obstructive event breaths. Finally, we repeated our ensemble average analysis, however utilizing non-AASM standard hypopnea scoring uniquely available in the MESA dataset, in which hypopnea events meet criteria of 30% to 50% airflow reduction but not associated with any oxygen desaturation events or arousal events. Unsurprisingly, this analysis shows an abatement of the initial observed difference in nadir ventilation (**Figure E4-C**, compare with Figure E4-A), further support that the events that terminate without an arousal contain a higher proportion of more severely obstructed events (and comparatively reduced ventilation) when compared to the events that terminate with an arousal.

Critically, while this observation affects the direct comparison of obstruction severity between arousal and non-arousal events, it does not compromise our primary findings regarding the relationship between arousal presence, intensity, and post-event ventilatory responses, as these analyses focus on the mechanistic effects of arousal characteristics rather than pre-event obstruction patterns.

Figure E4

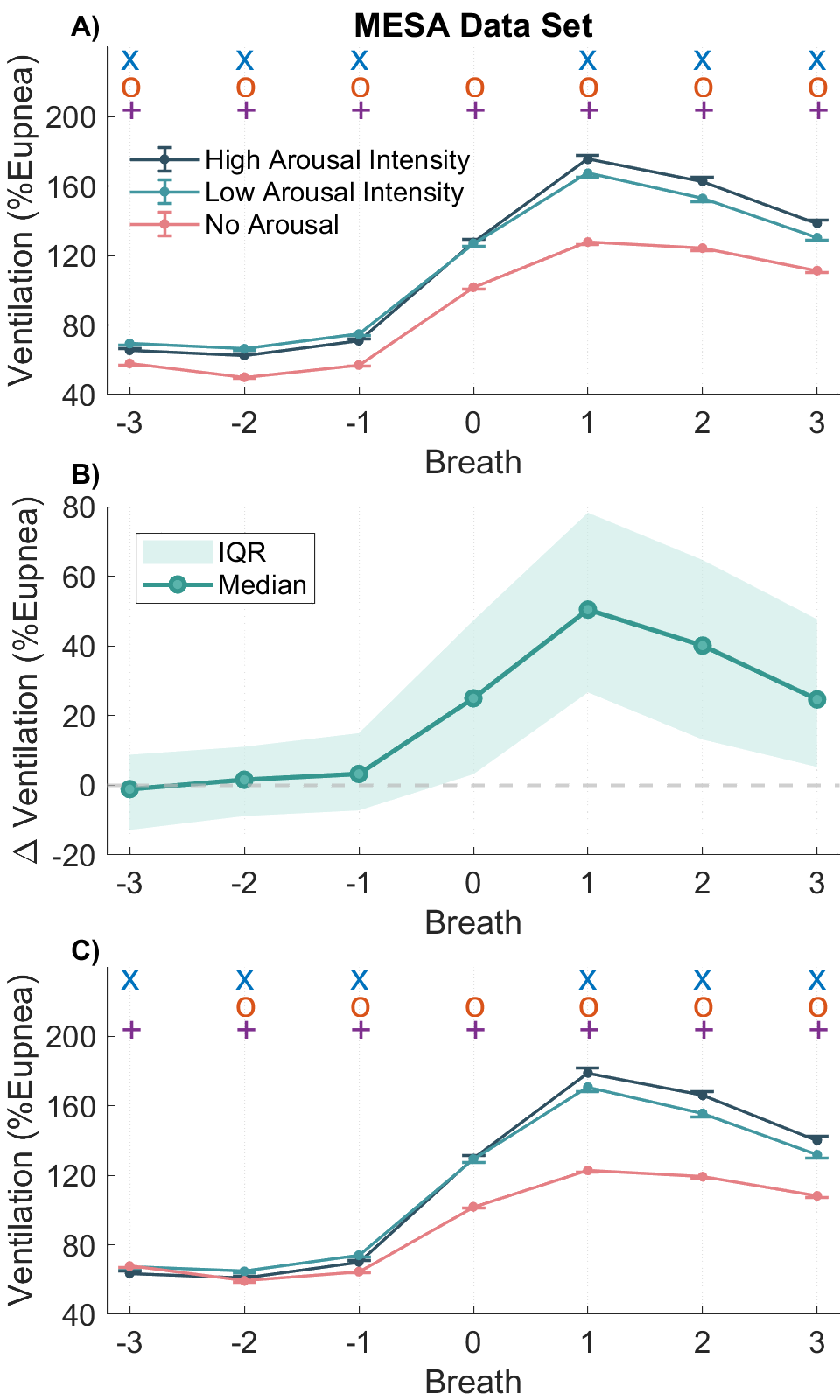

A): Original ensemble average analysis figure copied from main manuscript Figure 3A. B): Median and IQR of the within-participant ventilation difference between event terminated with arousal and event not terminated with arousal. C): Repeated ensemble average analysis of the ventilation for MESA Data Set with the inclusion of the additional scored hypopneas that are not associated with oxygen desaturation events or arousals.

### Age Stratification Analysis

Given the substantial age range differences between our two data sets (MESA: age range 54-94; Physiological Study: age range 21-74, see **Figure E5**), we conducted an age stratification analysis on MESA data set to examine whether arousal-mediated ventilatory responses vary across different age groups. This analysis addresses potential age-related confounding in our primary findings and explores the generalizability of arousal effects across different age groups.

We stratified the MESA participants into three age groups: age <65 years (n=686), 65≤age<75 years (n=571), and age ≥75 years (n=523). Physiological Study Data Set was not stratified due to smaller sample size. For each age stratum in the MESA Data Set, we applied our inter-event mixed-effects Model 1a, which allowed examination of age-specific arousal effects while maintaining our established analytical framework and controlling for event-specific ventilatory burden (**Figure E6**).

Age stratification revealed that *arousal intensity* shows a small increase in effect size with advancing age: 2.0%Eupnea per step increase vs. 2.2%Eupnea vs. 2.5%Eupnea (<65 years adults vs. 65-75 years adults vs. ≥75 years adults). If this age-related pattern was extrapolated to younger ages, it could partially explain why the Physiological Study Data Set demonstrated relatively small arousal intensity effects (0.3% eupnea per step, Model 1a) compared to MESA Data Set (showing 2.2% eupnea per step, Model 1a). This age-related increase may indicate that while younger adults may have similar overall arousal responses, older adults show stronger dose-dependent response relationships to intensity variations.

However, the *arousal presence* findings present a more complex picture. Here, age stratification reveals declining arousal presence effects with advancing age (35.2%Eupnea vs. 29.6%Eupnea vs. 24.6%Eupnea), suggesting younger adults should show stronger responses. This pattern appears inconsistent with the Physiological Study Data Set’s relatively modest arousal presence effect (22.8%Eupnea, Model 1a in the main analysis) compared to MESA Data Set (30.5% eupnea), given Physiological Study Data Set’s younger population. While the invasive instrumentation in the Physiological Study Data Set may play a more substantial role in determining arousal presence effects, the discrepancy observed in this analysis suggests that factors beyond age distribution alone influence the effects of arousal presence.

These findings highlight the complexity of arousal-ventilation relationships across different study populations and suggest that age represents only one important factor among several that influence these physiological responses. While age helps explain arousal intensity differences between datasets, the arousal presence patterns indicate additional unmeasured factors warrant consideration in future multi-cohort analyses of sleep-disordered breathing physiology.

Figure E5

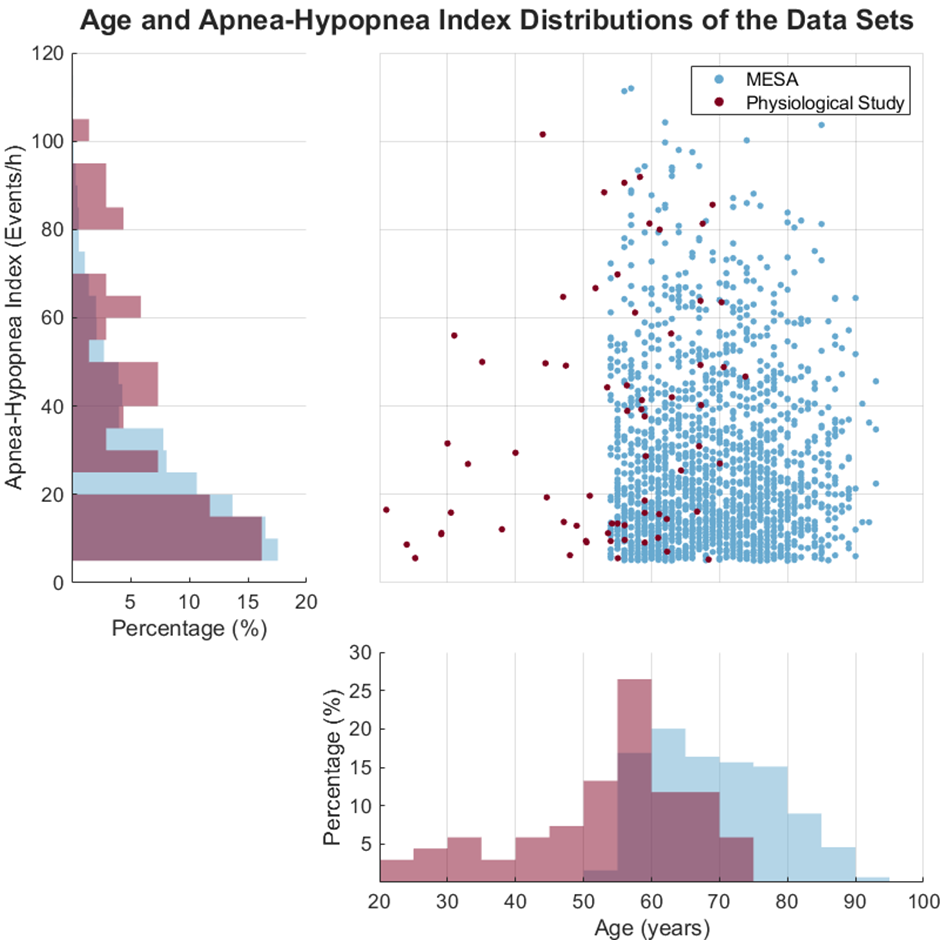

Scatter plot and histograms showing the age and OSA severity (Apnea-Hypopnea Index) differences and distributions in MESA (shown in blue) and the Physiological Study Data Set (shown in red).

Figure E6

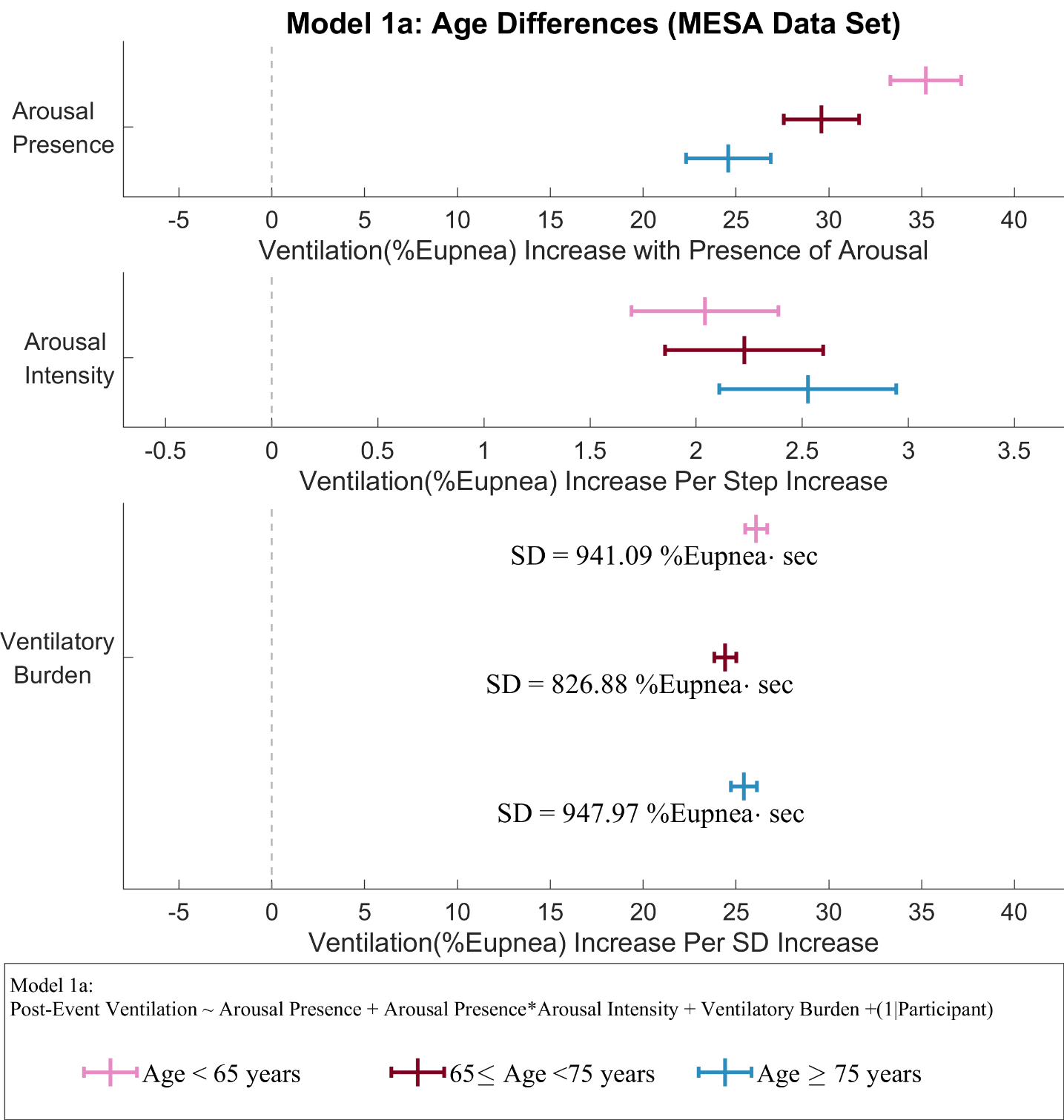

Forest plot shows the repeated inter-event analysis using Model 1a where participants were stratified by the age groups in the MESA Data Set. Each color represents each stratum. The center vertical bar represents the coefficient estimate and the horizontal error bar with end caps represents the 95% CIs.

### Flow Limitation at Event Termination

The severity of the flow limitation (i.e. upper airway conductance) was calculated using a previously validated machine-learning model (1). The degree of flow limitation was quantified throughout the night on a breath-by-breath basis as the ratio of airflow and ventilation effort (*i.e.*, Flow-Drive ratio or Flow:Drive).

Within each subject, all the scored obstructive events were aligned at the event-termination breath (Breath 0) and then stratified into three categories: high-intensity event vs. low-intensity event, and non-arousal event. Ensemble average flow-limitation were calculated for each category and each individual, identical to the main manuscript. The cohort level ensemble average was calculated by averaging all subject level averages. The result is shown in **Figure E7**. The arousal intensity was not associated with differences in flow limitation severity. However, the post-respiratory event breaths (breaths 1 and 2) for the respiratory events that were not terminated with an arousal are statistically significantly more flow-limited than the events accompanied by an arousal (mean flow-drive ratio for breaths 1 and 2, low-intensity vs non arousal: MESA, 0.79±0.10 vs. 0.72±0.10; Physiological Study 0.74±0.11 vs. 0.63±0.14).

Figure E7

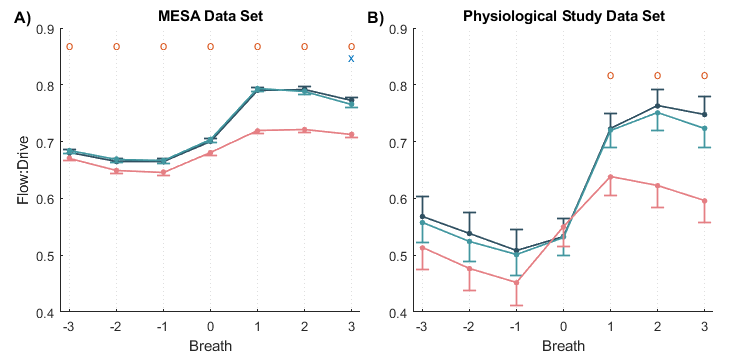

A) and B): Flow limitation (expressed as Flow:Drive) at the termination of respiratory events. “x”: p-value < 0.05 between high and low arousal intensity event at the respective breath; “o”: p-value < 0.05 between the event with arousal and those without arousal at the respective breath.

### Ensemble-Average of Ventilatory Drives

To further explore the temporal dynamics of arousal-induced hyperventilation, we conducted an ensemble-average analysis comparing model derived ventilatory drives (including $V_{chem}$ and VRA). Ventilatory drives were stratified on an event-basis for events terminated with high-intensity arousals, low-intensity arousal and non-arousal). Wilcoxon signed-rank tests were used to evaluate any statistically significant differences between 1) high-intensity vs low-intensity and 2) low-intensity vs no-arousal for each breath; and 3) high-intensity vs. no-arousal for each breath.

High-intensity arousal events were associated with statistically significant elevated post-event $V_{chem}$ comparing to low-intensity event at both Breath 1 (high- vs. low-intensity vs. non-arousal: 133.7±32.2 vs. 128.7±42.8 vs. 116.6±46.2 %eupnea) and Breath 2 (132.3±31.9 vs. 128.7±42.8 vs. 118.5±42.9 %eupnea) in MESA data set (**Figure E8-A**). The difference between low-intensity vs. non-arousal events and high-intensity vs. non-arousal events were also statistically significant. While the Physiological Study data set demonstrated similar trends (Breath 1: 148.3±33.9 vs. 140.9±31.3 vs. 130.1±46.2 %eupnea; Breath 2: 148.7±32.6 vs. 140.6±32.1 vs. 130.4±27.4 %eupnea; **Figure E8-B**), differences between high- vs. low-intensity events were not statistically significant. The difference between low-intensity vs. non-arousal events and high-intensity events vs. non-arousal events were statistically significant.

We repeated the ensemble average analysis for the Physiological Study data set using the ventilatory drive measured from the intra-esophageal diaphragmatic electromyograph (Edi) (**Figure E9**). This result confirmed the same pattern observed with PUP estimates: progressive reduction in ventilatory drive from high-intensity to low-intensity to non-arousal events in the post-event period (Breath 1: 161.3±13.5 vs. 131.0±4.3 vs. 116.7±5.4 %eupnea; Breath 2: 153.6±12.3 vs. 126.0±3.8 vs. 118.4±17.4 %Eupnea). Notably, the Edi measurements showed even greater separation between arousal intensity groups than PUP estimates, suggesting that direct neural drive measurements may be more sensitive to arousal intensity effects than computational approaches.

Figure E8

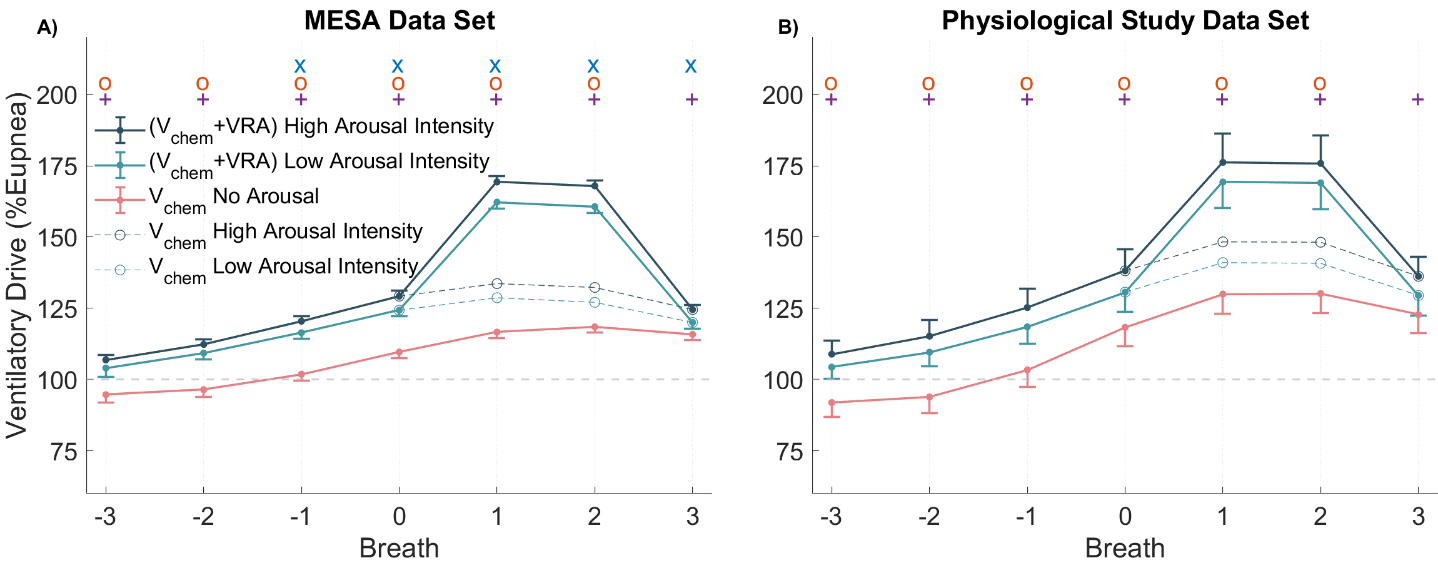

A) and B): Ensemble average plot showing the Vchem at the termination of respiratory event binned by High-, Low-intensity arousals and events not terminated with an arousal. VRA component is added to Breath 1 and 2 for high and low intensity events to accurately represent the total ventilatory drive. “x” represents p-value < 0.05 between high and low arousal intensity event for Vchem at the respective breath; “o” represents p-value < 0.05 between the event with low-intensity arousal and non-arousal for Vchem at the respective breath; “+” represents p-value < 0.05 between the event with high-intensity arousal and no-arousal for Vchem at the respective breath.

Figure E9

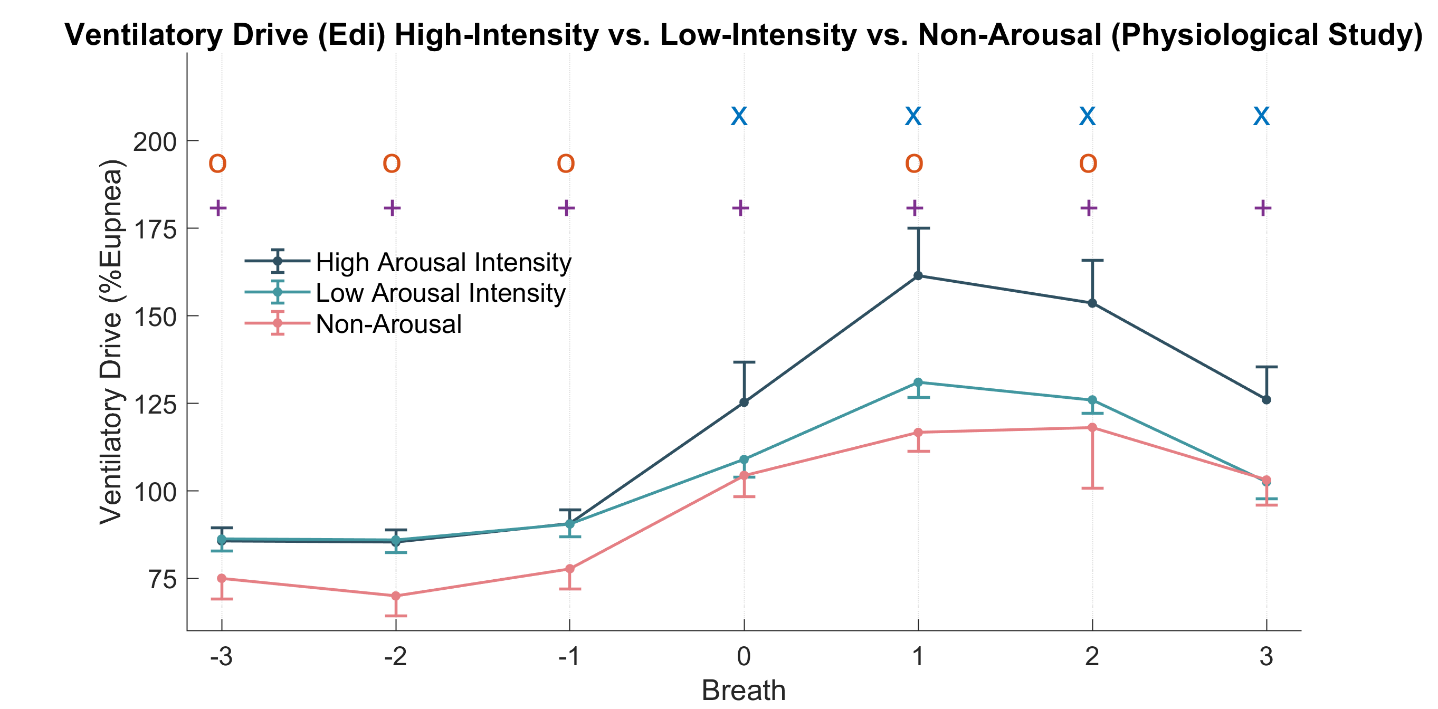

Ensemble average plot showing the ventilatory drive measured with intra-esophageal diaphragmatic electromyography (Edi) at the termination of respiratory event binned by High-, Low-intensity arousals and events not terminated with an arousal. “x” represents p-value < 0.05 between the event with high-intensity arousal and low-intensity for ventilatory drive at the respective breath; “o” represents p-value < 0.05 between the event with low-intensity arousal and non-arousal at the respective breath; “+” represents p-value < 0.05 between the event with high-intensity arousal and no-arousal at the respective breath.

### Association of Arousal Intensity with OSA Endo-Phenotypical Traits

In addition to the overnight average VRA, Each participant’s endo-phenotypical traits including one-minute cycle loop gain ($LG_{1})$, upper airway collapsibility ($V_{passive})$, upper airway muscle compensation ($V_{comp})$, and average arousal threshold $(ArTH)$ were computed as overnight aggregates using the PUP method formulated by Terrill et al. and Sands et al. (2-4). The $V_{passive}$, $V_{comp}$, and $ArTH$ magnitudes are normalized against the eupneic ventilation and presented as %eupnea. The cohort level statistics are summarized in **Table E1**.

Table E1: The summary of individual’s endo-phenotypical traits for both data sets.

| Characteristics | MESA | Physiological Study | |
| --- | --- | --- | --- |
| Mean $V_{passive}$(%Eupnea) $\boldsymbol{\pm}$SD* | 92.0 $\pm$ 11.3 | | 80.8 $\pm$ 24.7 |
| Mean ArTH (%Eupnea) $\boldsymbol{\pm}$SD | 124.9$\pm$ 25.6 | | 134.4 $\pm$ 38.9 |
| Mean $LG_{1}$ $\boldsymbol{\pm}$SD* | 0.52 $\pm$ 0.15 | | 0.66 $\pm$ 0.20 |
| Mean $V_{comp}$(%Eupnea) $\boldsymbol{\pm}$SD* | 5.5$\pm$ 17.4 | | -4.7 $\pm$ 23.1 |
| Definition of abbreviations:  Vpassive = Passive collapsibility; ArTH = Arousal Threshold; LG1 = one-minute cycle loop gain; Vcomp = Upper Airway Muscle Compensation  *a Statistically significant difference between two cohorts, it is not intentional in the study design. | | | |

Among the five calculated parameters, only overnight average VRA shows a statistical difference between high and low mean arousal intensity for patients in the MESA data set (**Figure E10)**. While a similar trend was observed in the Physiological Study data set, the difference was not statistically significant. Similarly, only VRA shows consistent positive association in both data sets and has a relatively large effect size (standardized $\beta=0.21$ for both data sets) comparing to the other endotypes, when these data were represented with arousal intensity treated as a continuous variable (**Figure E11**).

Figure E10

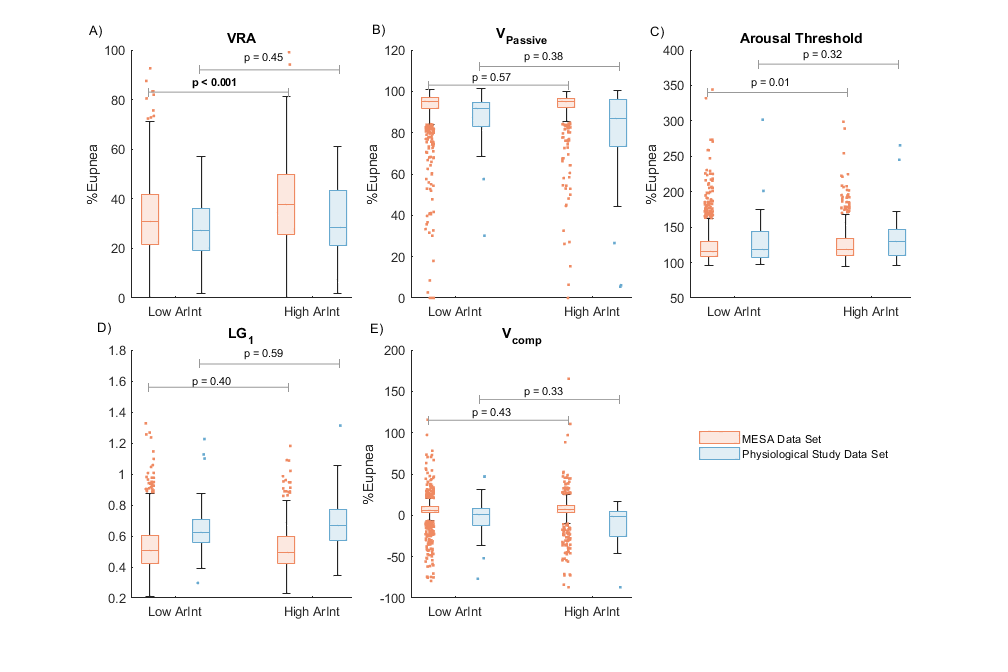

A)-E) Participants in each data set are separated according to the mean arousal intensity during NREM sleep. Low arousal intensity (Low ArInt) are individuals with mean arousal intensity < 5 and high arousal intensity (High ArInt) are individuals with mean arousal intensity $\geq$ 5. Each Individual’s endotypes, including overnight VRA, arousal threshold, $V_{\mathrm{Passive}}$, LG1 and $V_{\mathrm{comp}}$ are calculated and binned by high or low mean arousal intensity.

Figure E11

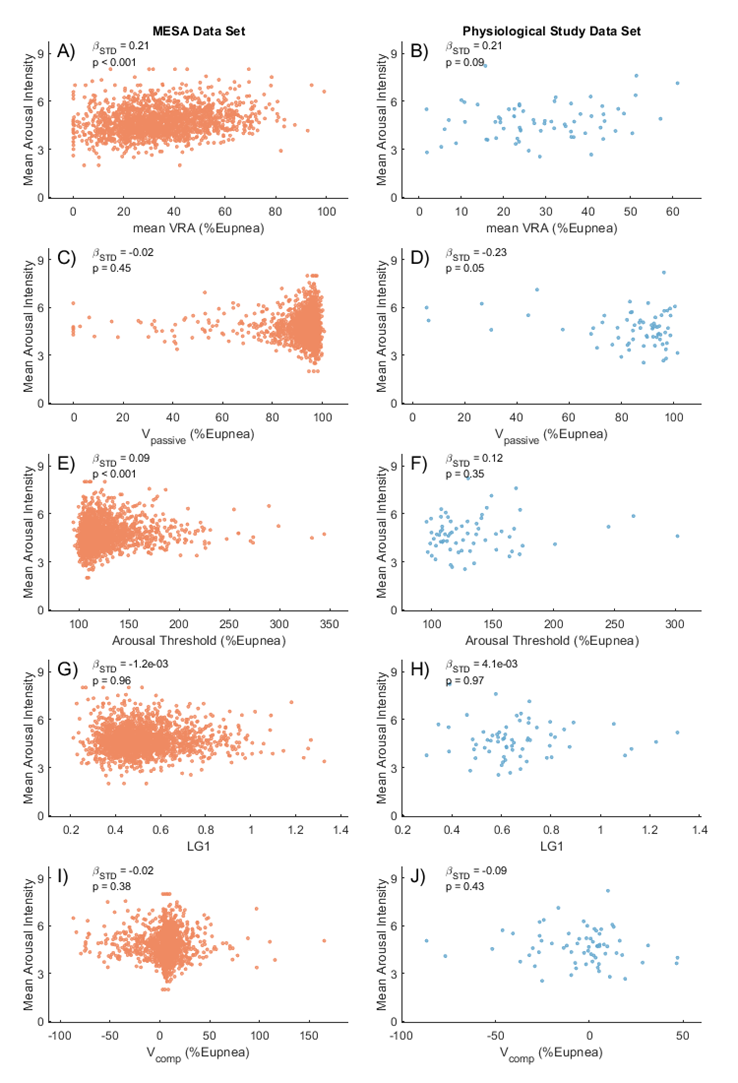

The mean arousal intensity of each participant is treated as a continuous variable and plotted against VRA (panel A and B), Passive anatomy (panel C and D), arousal threshold (panel E and F), one-minute cycle loop gain (panel G and H) and muscle compensation (panel I and J).

### Impact of REM Sleep Exclusion on Arousal-Ventilation Relationships

Our primary analysis excluded respiratory events occurring during REM sleep for two methodological reasons. First, the *PUP* method (2-4) used to estimate ventilatory drive has not been validated within REM sleep, where respiratory control mechanisms may differ substantially from NREM sleep. Since chemical drive estimates from *PUP* were integrated into Model 1 analyses, the inclusion of REM sleep could potentially introduce unexpected results. Second, potentially due to the additional instrumentation, the participants in the Physiological Data Set demonstrated markedly reduced REM sleep duration (Gell et al. (5)), and consequently, less events overall occur during REM sleep. Out of the included 67 participants, 30 did not have analyzable respiratory events in REM sleep. As such, the disproportionately low occurrence of REM sleep events may create an imbalanced representation across sleep stages.

However, to address concerns about REM exclusion, we repeated our primary inter-event analysis (Model 1, 1a and 1b) including all respiratory events regardless of sleep stage (**Figure E12**). This analysis incorporated 55211 additional REM events (including 16627 arousal and 38584 non-arousal events) from MESA data set and 740 (including 434 arousal and 306 non-arousal events) from the Physiological Study Data Set. The inclusion of these additional events had a modest impact on our results: the unadjusted model (Model 1) shows arousal presence increased post-event ventilation by 34.0%Eupnea in MESA Data Set (vs. 28.2% in NREM-only analysis) and 25.3% eupnea in the Physiological Study Data Set (vs. 20.6% in NREM-only). Arousal intensity effects were similar, in Model1, with each step increase in arousal intensity is associated with 4.7% eupnea ventilation increase in MESA Data Set (vs. 4.0% NREM-only) and 3.8% eupnea in the Physiological Data Set (vs. 3.3% NREM-only).

We conducted a comparison using MESA Data Set exclusively with the Model 1a which is adjusted with event-specific ventilatory burden, analyzing three conditions: NREM-only, REM-only, and combined REM+NREM (**Figure E13**). This comparison revealed notable sleep stage differences in arousal effects. In REM sleep, arousal presence produced a substantially larger ventilatory response (69.2%Eupnea increase) compared to NREM sleep (30.5%Eupnea increase), and each step increase in arousal intensity increase corresponded to 3.9% eupnea ventilation increase, compared to 2.2% eupnea per point in NREM sleep. This enhanced REM arousal effect likely reflects the physiological characteristics of REM sleep.

While the observed sleep stage differences, particularly the enhanced arousal presence effect in REM sleep, represent interesting physiological observations. These findings are consistent with previous studies reporting that arousal events in REM sleep produce greater responses than those in NREM sleep (6, 7). The mechanism which causes the difference in response in arousal between REM and NREM sleep is beyond the scope of the current study. Importantly, the observed difference does not alter our fundamental conclusions about arousal's contribution to post-event hyperventilation in NREM sleep. Future studies could examine the neurophysiological and respiratory control differences between REM and NREM sleep that contribute to varying arousal-ventilation relationships.

Figure E12

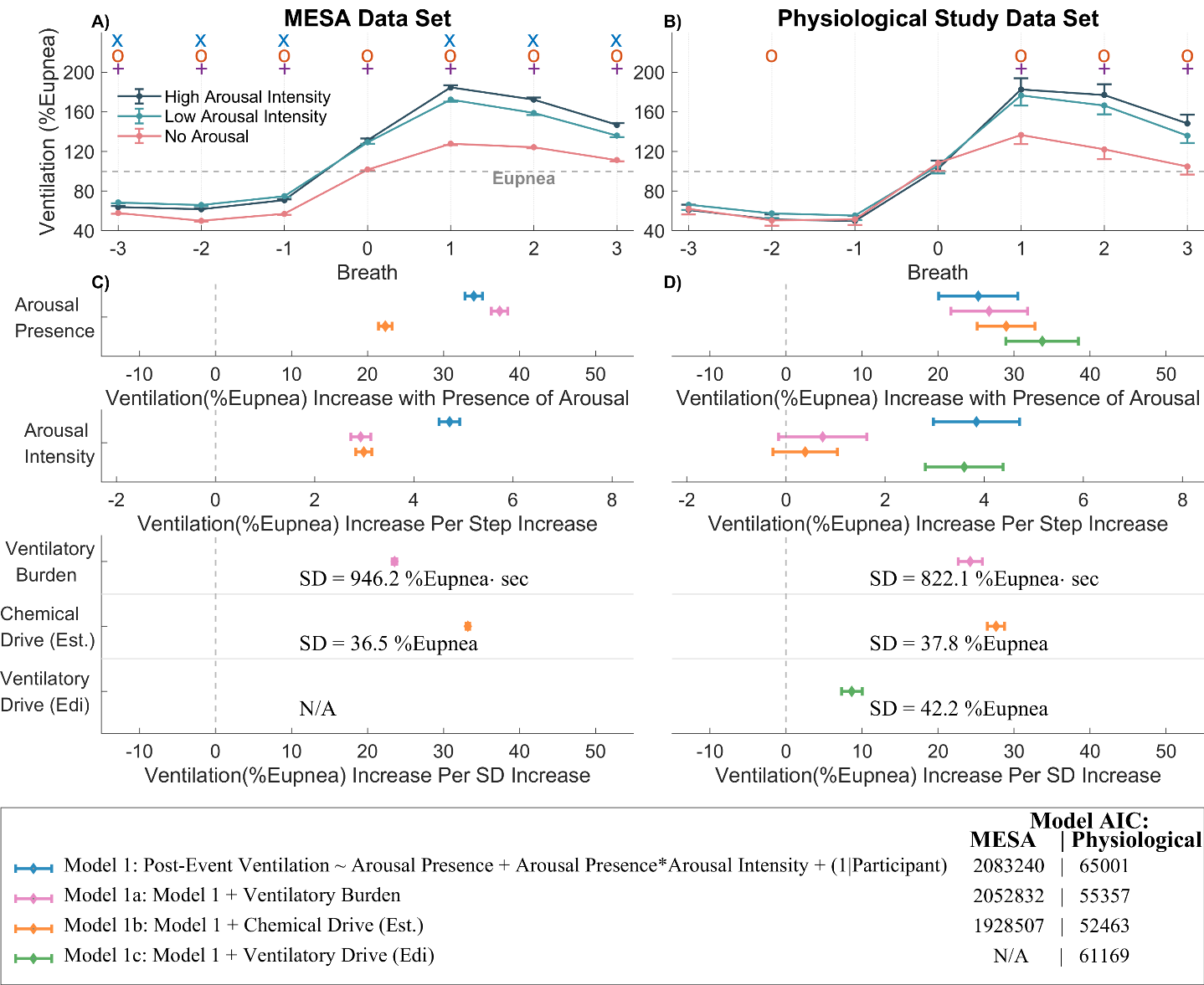

Repeated Inter-event analysis regardless of the sleep stage, to compare with Figure 3 which includes NREM sleep stage only.

Figure E13

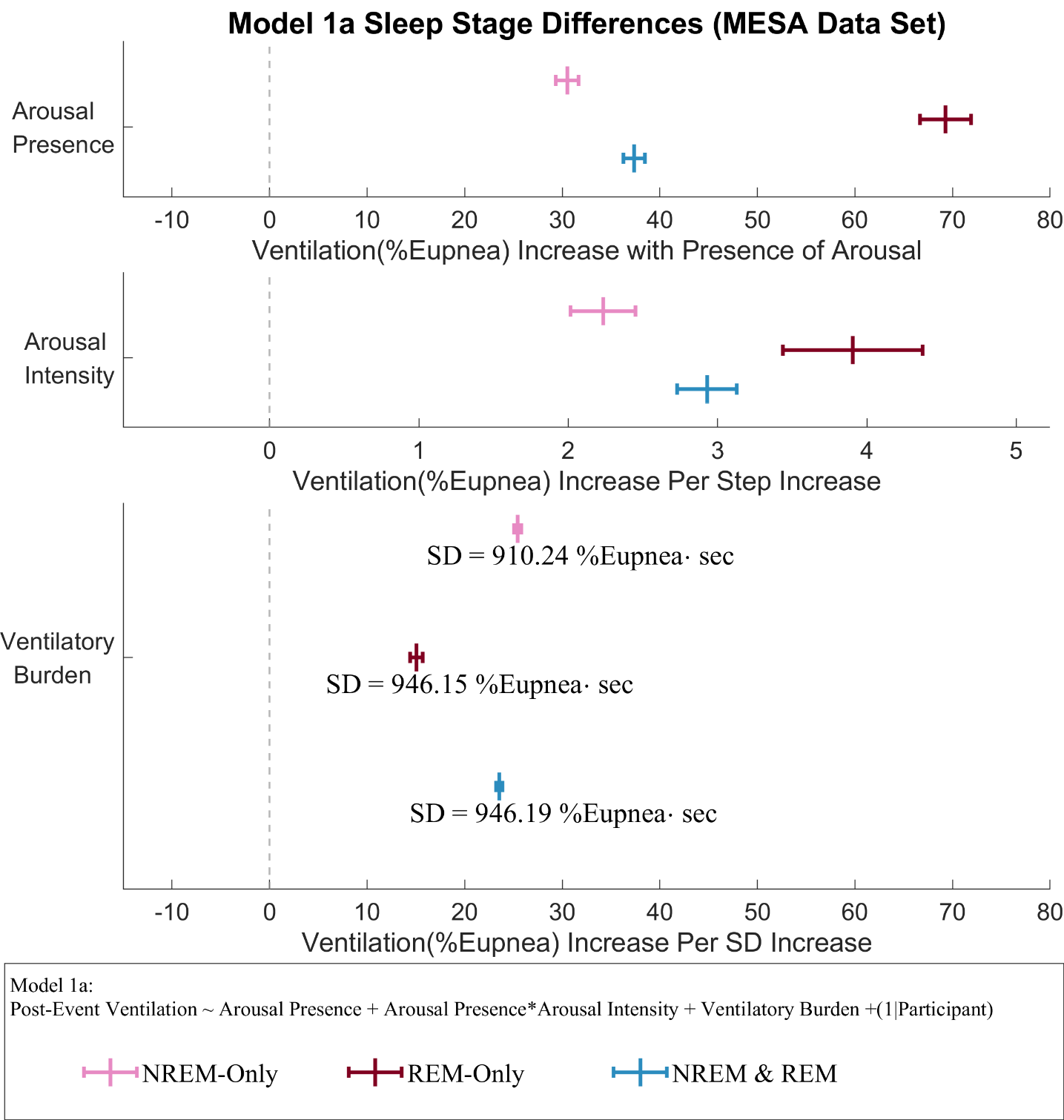

Forest plot showing the repeated inter-event analysis using Model 1a where respiratory events were stratified by the sleep stages in the MESA Data Set. Each color represents each stratum. The center vertical bar represents the coefficient estimate and the horizontal error bar with end caps represents the 95% CIs.
